# Prospective evaluation of accuracy of fine needle aspiration biopsy for breast lesions using the International Academy of Cytology Yokohama System for reporting breast cytopathology

**DOI:** 10.1101/2020.12.21.20248687

**Authors:** Shruti Agrawal, Michael Leonard Anthony, Pranoy Pal, Divya Singh, Anoushika Mehan, Ashok Singh, Prashant Joshi, Arvind Kumar, Bina Ravi, Shalinee Rao, Nilotpal Chowdhury

## Abstract

**Background:** Classification of breast leasions into different cytological groups (1-5) can accurately be done using the International Academy of Cytology (IAC) Yokohama System for reporting breast cytopathology. Fine needle aspiration biopsy (FNAB) of breast leasions has been considered to be the primary investigation in detecting breast cancers, especially in low-cost settings. The main objective of this study was to prospectively re-confirm the diagnostic accuracy of breast FNAB using the IAC Yokohama system. Additionally, separate separate secondary subgroup analysis was done to confirm the accuracy of breast FNAB excluding lymph-node positive and lymphadenopathy positive tumors.

**Material and methods:** A prospective study was done in a tertiary care centre in North India on patients undergoing core-needle /incisional/excisional biopsy of breast lesions between 1^st^ September 2019 to 30^th^ November,2020 (519 biopsies on 487 unique patients). Of these 519 histopathology biopsies, 505 had corresponding FNAB report of the same site. The FNAB was reported using the IAC Yokohama system and the most suitable category was allotted in every case. The rates of malignancy for each category and the accuracy of breast FNAB in diagnosing malignancy were calculated.

**Results:** Of the total 487 patients, 120 cases were benign on histology, while 367 were malignant. The rates of malignancy in benign, atypical, suspicious and malignant categories were 5%, 25%, 71% and 99.7% respectively. The diagnostic accuracy of atypical, suspicious and malignant categories was calculated as 90.1%, 95.2% and 93.3% respectively.

**Conclusion:** The high sensitivity and specificity for each scenario for all tumors and for each examined BIRADS category/ies suggest excellent accuracy for Breast FNAB using the IAC Yokohama system.

## Introduction

Breast Fine needle aspiration biopsy (FNAB) using the International Academy of Cytology (IAC) Yokohama System for reporting breast cytopathology can accurately classify breast lesions(1–5). It may be treated as a first line test in detecting breast cancers, especially in low-cost settings. Most of the studies on the accuracy of the IAC Yokohama system are on retrospective analysis(2,5,6). Also, many breast cancer patients are lymph node positive and easier to diagnose on FNAB due to the additional information gleaned from a lymph node imaging and FNAB. Likewise, it would be interesting to explore the accuracy of FNAB in lymph node negative cases. Keeping this in mind, we planned this study to prospectively re-confirm the diagnostic accuracy of breast FNAB using the IAC Yokohama system. We also planned separate secondary subgroup analysis to confirm the accuracy of breast FNAB using the IAC Yokohama system excluding lymph-node positive and lymphadenopathy positive tumors.

## Materials and Methods

This prospective study was conducted in a tertiary care teaching hospital in North India. Ethical approval for this study was obtained from the Institutional Ethical Committee.

All patients having breast symptoms in our centre are referred to an Intergrated Breast Care Centre (IBCC). Most of the patients are sysmptomatic who have been either self-referred, referred from outside by physicians/surgeons or referred from other departments from our institute. Only a small minority are screen detected. All the patients in the IBCC are examined clinically followed up by breast ultrasound and/or mammography. The BIRADS system is followed on breast imaging(7). Patients having BIRADS 4a lesions have ultrasound-guided (UG) breast FNAB, while those having BIRADS 4b, 4c, 5 lesions have both UG FNAB and UG core-needle biopsy. Additionally, patients having clinically suspicious lesions are sampled by core-needle biopsy regardless of the BIRADS score. At least 5 cores are normally attempted in each core needle biopsy. In patients having multiple malignant lesions, FNAB is done from all lesions, but core-needle biopsy may be performed only from the largest or most radiologically and clinically suspicious lesion/s. Non representative core-biopsies are followed by repeat core biopsies. In some cases, when the breast cancer involves the skin (stage IV a), FNAB from the primary lesion may be avoided and only a wedge biopsy or a punch biopsy taken. The FNAB is reported using the IAC Yokohama system in the Department of Pathology. The FNAB report ends by reporting the recommended categories (Inadequate, Benign, Atypical, suspicious of malignancy and Malignant). If the FNAB is reported as atypical, suspicious of malignancy or malignant in BIRADS 4a or lower cases, core needle biopsy is attempted on the next visit of the patient.

The present study was done on all patients having core-needle /incisional/excisional biopsy between 1^st^ September 2019 to 30^th^ November,2020 (519 biopsies on 487 unique patients). Of these 519 histopathology biopsies, 505 had corresponding FNAB report of the same site. Out of the 505 cases having cyto-histo correlation, eleven were further excluded from the analysis (six cases formed a part of an earlier study, five cases were inadequate/unrepresentative on histopathology). In a further seven cases having cyto-histological correlation, the Yokohama categories were not reported at the time of reporting, and were excluded from the primary analysis. This left 487 cases on which the primary analysis was performed. In all cases, the FNAB was reported before histopathology. The FNAB results were available if needed to the reporting histopathologist for the core-needle or excisional biopsies.

The IAC Yokohama category was deduced from the cytology report for the seven cases in which they were not a part of the original report. At the end of the study, supplementary sensitivity analysis was done including the patients who had a prospective Yokohama report but were excluded from the primary study (additional six cases), as well as including those who were not given a Yokohama category at the time of reporting (thus including seven cases). The characteristics of the patients not having cytological correlation or excluded from analysis are given in Supplementary table 1a to d.

**Table 1:** Showing the rates of malignancy with 95% Confidence Interval (C.I.) of the IAC Yokohama system categories for all the breast FNABs having cytological-histological correlation.

|  | Inadequate | Benign | Atypical | Suspicious for malignancy | Malignant |
| --- | --- | --- | --- | --- | --- |
| Non-malignant | 7 | 70 | 36 | 6 | 1 |
| Malignant | 3 | 4 | 12 | 15 | 333 |
| ROM | 0.3 | 0.05 | 0.25 | 0.71 | 0.997 |
| C.I. of ROM | [0.07, 0.65] | [0.01, 0.13] | [0.14, 0.40] | [0.48, 0.89] | [0.98, 1] |

For statistical analysis, all invasive carcinomas, DCIS, borderline and malignant Phyllodes tumors, sarcomas and lymphomas were considered as malignant. All other lesions (including benign Phyllodes tumors, atypical ductal hyperplasia, papilloma, fibroadenoma, fibrocystic change and acute/chronic inflammatory disease) were considered as benign. One case had a suspicious report on cytology, but the core-needle biopsy was non-representative with a requested repeat biopsy, and was excluded.

Statistical Analysis: The rates of malignancy (malignant cases/overall cases) with exact 95 % Confidence intervals were calculated for each category. The accuracy of breast FNAB in diagnosing malignancy was calculated for the following three diagnostic scenarios: i. Considering Only the benign category as non-malignant on cytology and the atypical, suspicious of malignancy and malignant categories as a malignant report; ii. Considering the benign and atypical categories as a non-malignant report and the suspicious of malignancy and malignant categories as a malignant report; and iii. Considering the benign, atypical and suspicious of malignancy categories as non-malignant, and only the malignant category as a malignant report. For all these three scenarios, the sensitivity, specificity, positive predictive value, negative predictive value with the 95% confidence intervals were calculated. This was followed by calculating the points of a Receiver Operating Characteristics (ROC) curve along with the area under the curve (AUC) and its bootstrap 95% confidence interval. Both empirical and binormal smoothed AUC were calculated.

The AUC was additionally calculated for the following additional analyses: i. Including the cases not given a Yokohama category at the time of reporting, but retrospectively given a category; ii. Including the cases which were already previously reported in an independent study along with the cases not given a Yokohama category at the time of reporting; iii. Including only cases which were lymph-node negative for malignancy along with the histopathological benign cases; iv. Including only patients who did not have any lymphadenopathy with malignancy along with the histopathological benign cases.

The R statistical environment along with the epiR (for sensitivity, specificity, PPV and NPV) and pROC (for ROC curve analysis) packages were used for statistical calculation(8,9).

## Results

The patients having histopathological correlation had a median age of 44 years (range: 15 to 85 years). The histopathological benign patients had a median age of 34 years (range: 15 to 75 years) while the histopathological malignant cases had a median age of 44 years (range: 15 to 85 years). There were 7, 94, 30, 26, 301 and 5 cases belonging to BIRADS 3, 4a, 4b, 4c, 5 and 6 categories respectively. One hundred and twenty cases were benign on histology, while 367 were malignant. Of the 120 benign cases, 15 had lymphadenopathy. Of the 367 histological malignant cases, 300 had lymphadenopathy out of which 221 showed metastasis. The rates of malignancy for the different IAC Yokohama categories for all the cases are given in Table 1. The sensitivity, specificity, PPV, NPV and diagnostic accuracy for each diagnostic scenario are given in Table 2. The AUC with 95 % Confidence intervals for the primary analysis, and all supplementary and subgroup analyses are given in Table 3. The histopathology diagnoses rendered for all cases are given in Table 4 grouped according to the observed cyto-histological correlation. The high sensitivity and specificity for each scenario for all tumors and for each examined BIRADS category/ies suggest excellent accuracy for Breast FNAB using the IAC Yokohama system.

**Table 2:** The sensitivity, specificity, positive predictive value (PPV), and negative predictive value (NPV) for breast FNAB using the IAC Yokohama system, under different diagnostic cut-offs for diagnosing malignant breast disease. The 95% Confidence intervals are given in square brackets.

|  | Atypical considered positive | Suspicious for malignancy considered positive | Malignant considered positive |
| --- | --- | --- | --- |
| Sensitivity | 0.989 [0.972 , 0.997] | 0.956 [0.93 , 0.975] | 0.915 [0.881 , 0.941] |
| Specificity | 0.619 [0.523 , 0.709] | 0.938 [0.877 , 0.975] | 0.991 [0.952 , 1] |
| PPV | 0.893 [0.859 , 0.922] | 0.980 [0.96 , 0.992] | 0.997 [0.983 , 1] |
| NPV | 0.946 [0.867 , 0.985] | 0.869 [0.796 , 0.923] | 0.783 [0.707 , 0.848] |
| Diagnostic Accuracy | 0.901 [0.871 , 0.927] | 0.952 [0.929 , 0.969] | 0.933 [0.907 , 0.954] |

**Table 3:** Showing the area under curve (AUC) of the ROC of FNAB reported by the IAC Yokohama system for diagnosing breast malignancy, with bootstrap 95% Confidence intervals (5000 bootstrap replicates for each analysis). The first row (bolded) shows the AUC of the primary analysis. Also analysed are the AUCs after adding the cases not reported by the IAC Yokohama system during the study period, after adding cases which were already reported in a separate study, after excluding lymph-node positive malignancies and after excluding malignant cases showing lymphadenopathy.

|  | Non-parametric AUC | Smoothed AUC |
| --- | --- | --- |
| **Overall** | **0.980 [0.965 , 0.991]** | **0.986 [0.976 , 0.994]** |
| Including cases not given a Yokohama category | 0.979 [0.965 , 0.99] | 0.986 [0.975 , 0.994] |
| Including cases not given a Yokohama category and cases excluded due to inclusion in a previous study | 0.979 [0.965 , 0.99] | 0.986 [0.975 , 0.993] |
| Excluding Lymph Node positive malignancies | 0.959 [0.934 , 0.98] | 0.968 [0.942 , 0.986] |
| Excluding lymphadenopathy positive malignancies | 0.945 [0.903 , 0.979] | 0.952 [0.902 , 0.982] |

**Table 4:**
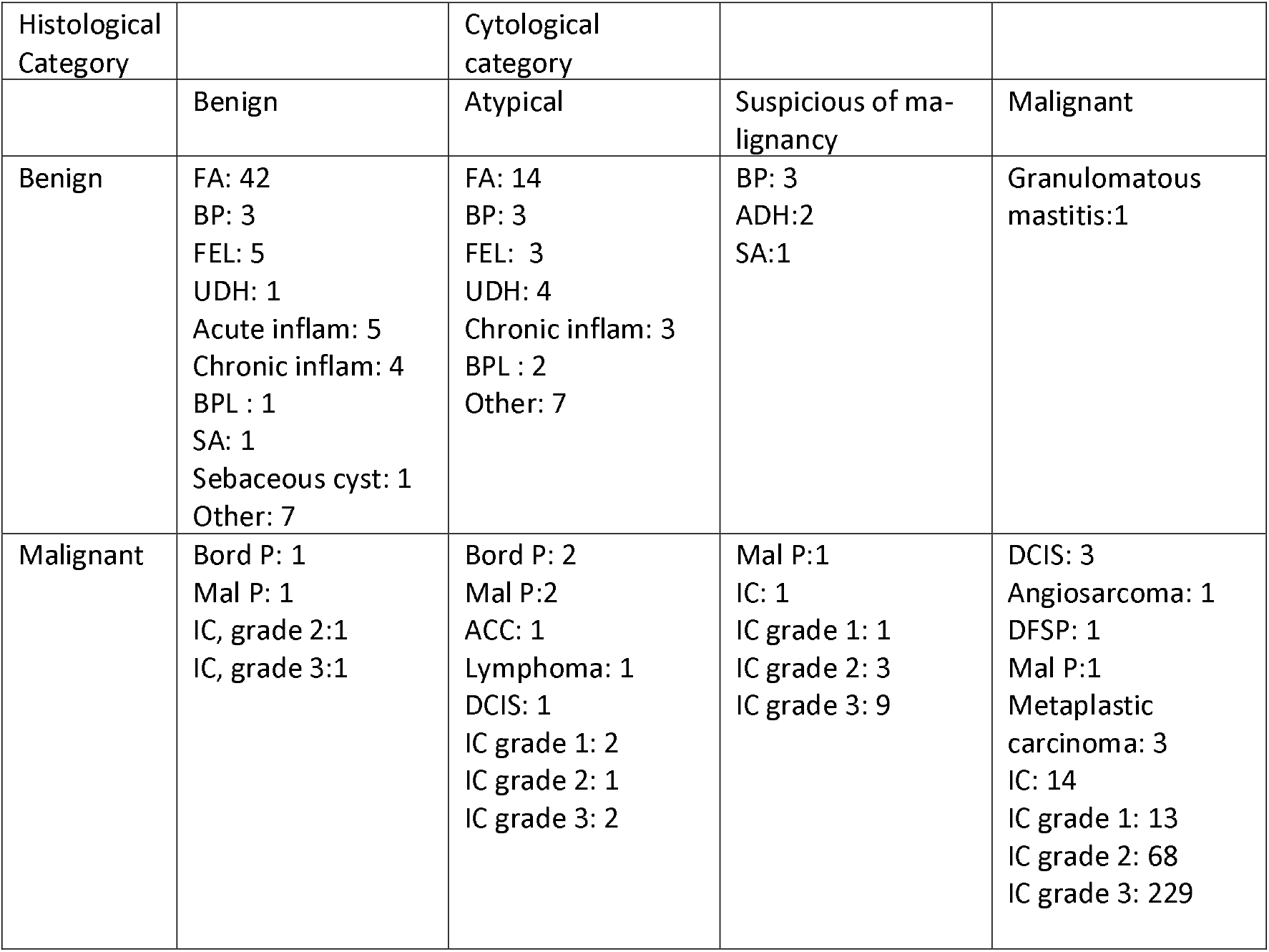
Showing the distribution of diagnosis in each cyto-histological category (FA: Fibroadenoma, BP: Benign Phyllodes tumor, FEL: Benign Fibrous/fibroepithelial lesion, UDH: Usual Ductal Hyperplasia, BPL: Benign Papillary Lesion, Acute inflam: acute inflammatory lesion, Chronic inflam: chronic inflammatory lesion, including granulomatous inflammation, Bord P: Borderline Phyllodes tumor, Mal P: Malignant Phyllodes Tumor, DCIS: Ductal Carcinoma-in-situ, IC: Invasive carcinoma, SA: Sclerosing adenosis, DFSP: Dermatofibrosarcoma Protuberans)

| Histological Category |  | Cytological category |  |  |
| --- | --- | --- | --- | --- |
|  | Benign | Atypical | Suspicious of malignancy | Malignant |
| Benign | FA: 42<br>BP: 3<br>FEL: 5<br>UDH: 1<br>Acute inflam: 5<br>Chronic inflam: 4<br>BPL : 1<br>SA: 1<br>Sebaceous cyst: 1<br>Other: 7 | FA: 14<br>BP: 3<br>FEL: 3<br>UDH: 4<br>Chronic inflam: 3<br>BPL : 2<br>Other: 7 | BP: 3<br>ADH:2<br>SA:1 | Granulomatous mastitis:1 |
| Malignant | Bord P: 1<br>Mal P: 1<br>IC, grade 2:1<br>IC, grade 3:1 | Bord P: 2<br>Mal P:2<br>ACC: 1<br>Lymphoma: 1<br>DCIS: 1<br>IC grade 1: 2<br>IC grade 2: 1<br>IC grade 3: 2 | Mal P:1<br>IC: 1<br>IC grade 1: 1<br>IC grade 2: 3<br>IC grade 3: 9 | DCIS: 3<br>Angiosarcoma: 1<br>DFSP: 1<br>Mal P:1<br>Metaplastic carcinoma: 3<br>IC: 14<br>IC grade 1: 13<br>IC grade 2: 68<br>IC grade 3: 229 |

## Discussion

Breast FNAB is an accurate diagnostic test with the added advantage of having a low cost and rapid reporting(10). The high accuracy of breast FNAB in diagnosing malignant disease confirms its suitability as a first-line diagnostic test. The ROM of the various diagnostic categories have been given and show significant differences from one another (Table 1).

The ROM of the benign category in the present study is probably higher (thus worse) than the actual ROM of the benign category. The FNAB cases with a benign FNAB diagnosis with discordant BIRADS score (4b, 4c, 5) or suspicious clinical feature were predominantly examined further by histopathology. The low-suspicion cases with a concordant benign FNAB diagnosis were not examined further, leaving out a large number of true negative cases. This probably brought down the reported specificity. Thus, the accuracy statistics reported is possibly biased downwards. Even then, the reported accuracy is on the high and comparable to other reported articles.

For the atypical and suspicious of malignancy categories, the confidence interval of the ROM includes the expected ROM of 15% and 85% respectively. However, the estimates of the ROM in the present paper and also a previous paper are closer to 25% for the atypical category and 70-75% for the suspicious category. In the present paper, a higher than expected number of Phyllodes tumors, especially borderline and malignant types, was partly responsible for the high reported ROM of the atypical category. Differentiating Borderline from benign Phyllodes tumor and benign Phyllodes tumor from cellular fibroadenoma on FNAB is known to be difficult, and may result in misclassification.

The ROM of the malignant category was satisfactory, but there was a false positive diagnosis of a Malignant Spindle cell neoplasm in what turned out to be granulomatous mastitis. This case was characterized by large spindled cells in a necrotic background admixed with acute and chronic inflammatory cells. The case was clinically as well as radiologically suspicious of malignancy (BIRADS score of 5: highly suggestive of malignancy). The necrosis on cytology was mistaken for the necrosis of malignancy, and large reactive epithelioid type histiocytes were mistaken for a spindle cell neoplasm.

For malignant cases, the diagnosis may have been made easier by the relatively higher stage of presentation of the patients. Therefore, subgroup analyses excluding lymph-node positive cancers and patients of cancers having lymphadenopathy were performed (Table 3). In both these scenarios, the diagnostic accuracy as estimated by the AUC were high and satisfactory.

To the best of our knowledge, this is the first prospective validation of the IAC Yokohama system. This strengths of this study are that this was a prospective study done on a large series having cyto-histological correlation for greater than 96% of the histopathology cases. Supplementary AUC curve analyses was were performed which included the small number of cases excluded from primary analysis (Table 3), and the diagnostic accuracy results were almost the same 0.1 % difference from the primary analysis for the empirical AUC.

The primary limitation that we faced in this study was the lack of information about the cases having FNAB benign diagnosis and having correlating clinico-radiological picture. The

nCOViD pandemic and resultant also resulted in getting a lower than expected histological correlation of the benign cases since the population was advised to largely avoid crowds and stay at home. The less serious patients as a result were deferred, with malignant cases being prioritized. We feel that this has resulted in a lower diagnostic accuracy statistic compared to the actual accuracy.

We also intended to carry out a study of the accuracy of the IAC Yokohama system in the various comparison with the BIRADS categories. The relatively low number of patients in some BIRADS categories prevented robust conclusion to be drawn from the present data. We intend to collect more data, and combine previous data to expand the time frame of data collection and carry out a separate analysis which will lead to hopefully robust statistical conclusion about the accuracy of FNAB in different radiological risk categories.

In conclusion, this study re-confirms the excellent accuracy of breast FNAB using the IAC Yokohama system in diagnosing breast malignancies. Furthermore, we document for the first time its suitability in BIRADS 4a and 4b lesions corresponding to low and moderately suspicious breast lesions. Breast FNAB should retain its relevance in situations where cost and time are limited resources.

## Data Availability

All the data used in the study has been provided in the manjscript and as supplementary files.

## References

1. Hoda RS, Brachtel EF. International Academy of Cytology Yokohama System for Reporting Breast Fine-Needle Aspiration Biopsy Cytopathology: A Review of Predictive Values and Risks of Malignancy. Acta Cytol [Internet]. 2019 [cited 2019 Oct 24];63[Suppl. 4):292–301. Available from: http://www.ncbi.nlm.nih.gov/pubmed/31141809

2. Montezuma D, Malheiros D, Schmitt FC. Breast Fine Needle Aspiration Biopsy Cytology Using the Newly Proposed IAC Yokohama System for Reporting Breast Cytopathology: The Experience of a Single Institution. Acta Cytol [Internet]. 2019 Feb 15 [cited 2019 Oct 24];63[Suppl. 4):274–9. Available from: http://www.ncbi.nlm.nih.gov/pubmed/30783035

3. Wong S, Rickard M, Earls P, Arnold L, Bako B, Field AS. The International Academy of Cytology Yokohama System for Reporting Breast Fine Needle Aspiration Biopsy Cytopathology: A Single Institutional Retrospective Study of the Application of the System Categories and the Impact of Rapid Onsite Evaluation. Acta Cytol [Internet]. 2019 Jun 1 [cited 2019 Oct 23];63[Suppl. 4):280–91. Available from: https://www.karger.com/Article/FullText/500191

4. McHugh KE, Bird P, Sturgis CD. Concordance of breast fine needle aspiration cytology interpretation with subsequent surgical pathology: An 18LJyear review from a single subLJSaharan African institution. Cytopathology [Internet]. 2019 Sep 22 [cited 2020 Aug 21];30[5):519–25. Available from: https://onlinelibrary.wiley.com/doi/abs/10.1111/cyt.12696

5. Agarwal A, Singh D, Mehan A, Paul P, Puri N, Gupta P, et al. Accuracy of the International Academy of Cytology Yokohama system of breast cytology reporting for fine needle aspiration biopsy of the breast in a dedicated breast care setting. Diagn Cytopathol [Internet]. 2020 Oct 5 [cited 2020 Oct 23];dc.24632. Available from: https://onlinelibrary.wiley.com/doi/10.1002/dc.24632

6. De Rosa F, Migliatico I, Vigliar E, Salatiello M, Pisapia P, Iaccarino A, et al. The continuing role of breast fineLJneedle aspiration biopsy after the introduction of the IAC Yokohama System For Reporting Breast Fine Needle Aspiration Biopsy Cytopathology. Diagn Cytopathol [Internet]. 2020 Aug 4 [cited 2020 Aug 21];dc.24559. Available from: https://onlinelibrary.wiley.com/doi/abs/10.1002/dc.24559

7. D’Orsi CJ, Sickles E, Mendelson E, Morris E. ACR BI-RADS Atlas: Breast Imaging Reporting and Data System; Mammography, Ultrasound, Magnetic Resonance Imaging, Follow-up and Outcome Monitoring, Data Dictionary. 5th ed. Reston, Virginia: ACR, American College of Radiology; 2013.

8. R Core Team. R: A Language and Environment for Statistical Computing [Internet]. Vienna, Austria: R Foundation for Statistical Computing; 2019. Available from: http://www.r-project.org/

9. Robin X, Turck N, Hainard A, Tiberti N, Lisacek F, Sanchez J-C, et al. pROC: an open-source package for R and S+ to analyze and compare ROC curves. BMC Bioinformatics. 2011;12:77.

10. Field AS, Raymond WA, Rickard M, Arnold L, Brachtel EF, Chaiwun B, et al. The International Academy of Cytology Yokohama System for Reporting Breast Fine-Needle Aspiration Biopsy Cytopathology. Acta Cytol [Internet]. 2019 [cited 2019 Oct 24];63[Suppl. 4):257–73. Available from: http://www.ncbi.nlm.nih.gov/pubmed/31112942

